# Associations Between Social Responsiveness and Sleep disturbance are Modulated by Chronotype in Early Adolescence: Cross-Sectional and Prospective Findings from 10,108 Participants of the Adolescent Brain and Cognitive Development (ABCD) Study

**DOI:** 10.64898/2026.06.20.26356092

**Authors:** Cathy Wyse, Mailton Vasconcelos, AungMyat Phyo, Enya Nordon, Lorna M. Lopez

## Abstract

**Study Objectives:** Sleep disturbance is prevalent in people with neurodevelopmental disorders such as autism, but is not clear whether it occurs as an endophenotype or secondary to other behaviours. The ABCD Study is a population-based longitudinal study that monitors the health, demography and lifestyle of over 11,000 children in the US. In this study we leverage these data to investigate whether traits consistent with autism (social responsiveness) are associated with sleep disturbance independent of lifestyle and other behavioural measures.

**Methods:** Autistic traits were assessed using the Social Responsiveness Scale at age 11, and sleep disturbance and behavioural outcomes were assessed at ages 11 and 13 years using the Sleep Disturbance Scale, and the Child Behaviour Check List, respectively. Demographic, health and lifestyle-related variables were assessed by caregiver questionnaires. Regression models were applied to investigate associations between autistic traits and sleep outcomes.

**Results:** There was a significant cross-sectional association between sleep disturbance and SRS at age 11 years old that was independent of sex, ethnicity, socioeconomic position, physical activity, sedentary behaviour and anxiety/depression (β = 0.12, 95% CI (0.07, 0.17); p < 0.001), that persisted at age 13, and that was modulated by chronotype, with evening types showing a stronger association.

**Conclusions:** Social responsiveness assessed in early adolescence (age 11) were associated with sleep disturbance independent of multiple confounding factors and were prospectively associated with sleep disturbance at age 13 years. These findings contribute to the evidence that disruption of sleep and circadian timing may have a primary role in the mechanisms that mediate autistic traits.

**Statement of significance:** Sleep disturbance is prevalent in people with neurodevelopmental conditions including autism and is associated with the severity of social, behavioural, and sensory symptoms. Evidence from studies of high-risk infants and animal models suggest that sleep problems emerge early in development, and may reflect underlying neurobiological vulnerability rather than secondary effects of other aspects of neurodevelopmental conditions. This study provides population-level evidence that autistic traits are independently and longitudinally associated with sleep disturbance across early adolescence. This relationship was moderated by chronotype, was stronger in females, and clustered within families, implicating shared circadian or genetic mechanisms. Future research should focus on prospective and mechanistic studies to assess the aetiological significance of sleep disturbance in autism.

## Introduction

Sleep problems are a common feature of neurodevelopmental conditions in children^1–3^ with significant consequences for their quality of life and that of their siblings and parents. Much of the current evidence supporting these associations is derived from case-control studies in clinical populations of children with a diagnosis of autism and are often confounded by comorbid conditions that affect sleep (e.g. ADHD, epilepsy, intellectual disability)^4^ and also by the use of sleep-modifying medication by many autistic children.^5^ Furthermore, assessment of sleep by caregivers of autistic children can be biased by their own sleep disturbance or by the child’s capacity to communicate, while objective methods of sleep assessment (actigraphy, polysomnography) are often poorly tolerated. The small sample sizes and wide age ranges of many previous studies cannot account for the developmental changes in sleep and circadian timing that occur across adolescence in both neurotypical and children with neurodevelopmental conditions (delayed circadian phase and reduced sleep duration).^6,7^ Large sample sizes are required to account for the wide variability in lifestyle and demographic factors, pubertal stage and comorbid psychopathology that could confound detection of effects on sleep that are a primary correlate of autism and other neurodevelopmental conditions.

Longitudinal, population-based studies can capture the trajectory of sleep development in early life and provide sufficient statistical power and detailed measurement of covariates to investigate whether changes in sleep among children with neurodevelopmental conditions are independent of behavioural comorbidities and other confounding factors. Studies in population-based cohorts have already reported associations between autism or autistic traits, and sleep disturbance that are consistent with the findings of case-control studies.^8–10^ Disruption of circadian and sleep timing are apparent in early infancy in autistic children^11–13^ and are associated with the presence and severity of symptoms including social affect, sensory difficulties and restrictive behaviours,^1,14^ a close association that has led to the hypothesis that sleep and circadian disruption is of aetiological significance in autism development.^15–18^

Circadian rhythms are generated by a molecular timing mechanism present in all nucleated cells that confers 24h periodicity on physiology and behaviour. Chronotype refers to daily preference for evening or morning activity, while social jetlag is a term taken to indicate misalignment between endogenous circadian timing and the timing of social (school or work) cycles. Circadian rhythms are disrupted in autistic children^19^ and a core component of the circadian timing mechanism (melatonin secretion) is an effective point of intervention.^20,21^ Experimental disruption of circadian rhythms initiated in juvenile (P21),^22^ and adolescent (P21-42)^19^ mice induced behaviours consistent with autism (diminished sociability, reduced novelty preference and repetitive behaviour), while mice carrying mutations in candidate autism genes (*Shank3*; *Scn2a*) have downregulated clock gene expression, and disrupted sleep and behavioural rhythms.^18,23–25^ Sleep disturbance profoundly affected neurodevelopment and the emergence of autistic-like behaviours in animal models in early life but not in adulthood,^24,26^ and significantly, these behaviours were rescued by manipulation of sleep,^27^ suggesting that sleep disturbance in early life could be a risk factor, and a novel point of intervention in autism.

Behavioural findings in animal models may not represent the human autistic phenotype but genetic studies in humans provide strong support for the underlying conclusions. For example, genetic correlation^28,29^ and Mendelian randomisation studies^28^ in *UK Biobank* and the *FinnGen Study* support a causal role for disruption of sleep and circadian timing mechanisms in autism, and people with rare variants in a gene encoding a core component of the circadian timing mechanism (*BMAL1*) have an autistic phenotype.^30^ This genetic evidence of causal associations in studies of autistic people in addition to the induction of autistic-behaviours by sleep and circadian disruption in animal models support phenotypic evidence that disruption of circadian and sleep timing is not just a consequence of autism, but an endophenotype embedded in its polygenic genetic architecture.

In this study, we leverage the large sample size and longitudinal study design of the largest population-based study of adolescent neurodevelopment to date (ABCD Study^31^) to investigate cross sectional and prospective associations between traits consistent with autism (social responsiveness) and sleep disturbance across adolescence. We use the Social Responsiveness Scale to assess traits associated with autism, given that autistic traits occur as a continuum across the population. The aim of this study is to assess whether sleep disturbance, social jetlag and chronotype are associated with autistic traits in a large population-based cohort study, and if any relationship persists through adolescence and is independent of demographic, lifestyle and comorbid behavioural conditions that could confound detection of a primary association of sleep disturbance and autism.

## Methods

### Participants

The study sample were participants of the ABCD Study, a longitudinal study of brain development across adolescence with school-based recruitment of 11,875 children aged 9-10 from 21 sites across the US.^32^ Further details about the ABCD Study can be found at http://abcdstudy.org. Centralised institutional ethical review board approval was obtained from the University of California, San Diego (IRB# 160091). Study site approvals were obtained from their respective institutional review boards. The caregivers of participants provided written informed consent, and the child participants provided written assent. The measures included caregiver completed questionnaires, interviews, neurocognitive interviews and neuroimaging.^31^ Details of all outcome, exposure and co-variables included in this study are given in Tables S1 and S2 and the timing of follow-up assessments are shown in Table S2. Since not all measures were collected at each follow-up appointment, for the purposes of this study, year 1 (aged 10-11, early adolescence) was taken as timepoint 1, and year 3 (aged 12-13, mid-adolescence) as the follow-up time point. Measures that were not available at either of these timepoints were imputed from the nearest available data (details of imputed data are given in Table S3). In other words, where data were not collected during timepoint 1 or 2 in the ABCD Study, we inputted data from the nearest available year that that specific information was available.

### Demographic and Lifestyle Data

Information on demographics (child sex, ethnicity, and total household income) was given by the child’s main caregiver. Ethnicity was reported by the caregiver and categorised as "White", "Black", "Asian" or "Other". Screen time on a typical day was reported by the caregiver as time on a computer, cell phone, TV, tablet, or other electronic device, and included both weekday and weekend screen time. Socioeconomic position (SEP) was calculated from the residential area deprivation index [kind], higher values indicate higher levels of deprivation. Physical activity was reported by the child as the number of days the child was physically active (> 1h per day) in the last week. Physical activity was assessed using the ABCD Youth Risk Behavior Survey item, which asks the children to report the number of days during the previous week on which they were physically active for at least 60 minutes, yielding a score from 0 to 7 days.

### Sleep Disturbance

The Sleep Disorders Scale for Children (SDSC) was a caregiver-completed questionnaire to assess sleep disorders in children over the last 6 months. The SDSC is a 26-item inventory that consists of six sleep disorder subscales: (i) disorders of arousal or nightmares, (ii) disorders of initiating and maintaining sleep, (iii) disorders of excessive somnolence, (iv) sleep breathing disorders, (v) sleep hyperhidrosis, and (vi) sleep–wake transition disorders. The sum of the subscales (total score) ranges from 26 to 130 points with higher scores representing lower sleep quality, and total score ≤ 39 indicating normal sleep.^33^ The SDSC includes an item on the child’s hours of nightly sleep that was used to indication sleep duration.

### Cognition

Cognitive ability was assessed using the NIH Toolbox cognition measures (http://www.nihtoolbox.org) that generates a total cognitive score based on seven tasks that assesses episodic memory, executive function, attention, working memory, processing speed, and language abilities.^34^

### Traits Consistent with Autism

The Social Responsiveness Scale (SRS) is a caregiver completed questionnaire with 11 items that assess social cognition, communication and mannerisms with higher scores indicating greater severity of social impairment. The SRS has had widespread use for quantitative assessment of autistic social impairment in research and public health settings, with over 20 official foreign language translations. The SRS cannot diagnose autism, but is a useful tool for continuous quantitative measure of traits associated with autism that has been validated against autism diagnoses derived from the Autism Diagnostic Interview-Revised (Constantino et al., 2003). The SRS describes core features of the autistic phenotype including receptive, cognitive, expressive, and motivational aspects of social behaviour. It includes three DSM-IV autism domains; reciprocal social behaviour (e.g. “*Has difficulty making friends, even when trying his or her best*”), stereotyped and repetitive behaviours (e.g. “*Has more difficulty than other children with changes in his or her routine*”), and communication impairments (e.g. “*Has trouble keeping up with the flow of normal conversation*”).^35,36^ There is no reliable information on autistic diagnoses in the ABCD Study, and the scores from the SRS are a measure of the continuum of autistic traits across a sample of the US population.

### Behavioural Symptoms

The Child Behaviour Checklist (CBCL) is a caregiver-completed questionnaire to assess emotional behaviour over the prior 6 months.^37^ The CBCL in the ABCD Study has 112 different items that each caregiver answered about their child’s behaviour (e.g., “*Show little interest in things around him/her*”) using a 3-point Likert-type scale (0 = Not True, 1 = Somewhat or Sometimes True, 2 = Very True or Often True). These answers are then used to calculate summary scores of internalizing and externalizing behaviours, with higher scores indicating more emotional, behavioural, and social problems. Externalising and internalising symptoms were included to assess the impact of co-occurring psychopathology.

### Chronotype and Social Jetlag

Parameters describing chronotype and social jetlag defined and derived in the ABCD Study were used in this study. The Youth Munich Chronotype Questionnaire (MCTQ) is a 17-item questionnaire that assesses daily preference (chronotype) in terms of the midpoint between sleep onset and offset on free days, with scores ranging from 16 to 40.^38^ The ABCD Study defined social jetlag as the absolute difference between mid-sleep time on free days and mid-sleep time on school/work days with higher values reflecting a greater degree of misalignment between biological and social circadian timing. Chronotype was treated as a continuous variable (and categorized into morning/intermediate/evening types), and social jetlag was modelled as a continuous variable.

### Data Analysis

Participant characteristics were summarised using mean and 95% confidence intervals for continuous variables, and frequency and percentage for categorical variables. Differences in covariables across the categories of the outcome variables (sleep disturbance, social jetlag) were assessed by χ2-test for categorical variables and univariate regression modelling for continuous variables (Tables 1, 2). A series of linear mixed-effects models were fitted to examine the association between social responsiveness and social jetlag, and sleep disturbance during adolescence (Tables S5 and S6). The dependent variable was sleep disturbance or absolute social jetlag (calculated as the absolute difference in mid-sleep timing between free and school nights). The social responsiveness scale was centred to give a continuous, standardized total score that could be applied to test interactions with other covariables. All models included random intercepts at an individual level nested within families to account for within-subject correlation across timepoints and non-independence among siblings or twins.

**Table 1.** Demographic, sleep and psychosocial summary data of sample of participants of the ABCD Study used in this study at 1-year follow up wave compared to 3-year follow up. These data illustrate the changes in each variable between ages 11 – 13 years.

|  | Age 11 years old<br>(N=11182) | Age 13 years old<br>(N=9840) | p |
| --- | --- | --- | --- |
| **Sex** |  |  |  |
| male | 5824 (52 %) |  |  |
| female | 5352 (48 %) |  |  |
| Missing | 6 (0.1%) |  |  |
| **Age (months)** |  |  |  |
| mean (95% CI) | 131 (130, 131) | 154 (154, 155) |  |
| Missing | 1 (0.0%) | 1 (0.0%) |  |
| **Puberty** |  |  |  |
| early puberty | 1790 (16 %) |  |  |
| mid puberty | 2461 (22 %) |  |  |
| late puberty | 1926 (17 %) |  |  |
| post puberty | 1115 (10 %) |  |  |
| Missing | 3890 (34.8%) |  |  |
| **Ethnicity** |  |  |  |
| White | 7842 (70 %) |  |  |
| Black | 1609 (14 %) |  |  |
| Asian | 391 (3 %) |  |  |
| Other/Mixed | 1086 (10 %) |  |  |
| Missing | 254 (2.3%) |  |  |
| **SEP** |  |  |  |
| mean (95% CI) | 39.66 (39.15, 40.18) | 38.82 (38.28, 39.36) |  |
| Missing | 795 (7.1%) | 661 (6.7%) |  |
| **Sleep Disruption** |  |  |  |
| mean (95% CI) | 36.65 (36.5, 36.8) | 36.01 (35.86, 36.17) | <0.001 |
| **Sleep Duration** |  |  |  |
|  |  |  | <0.001 |
| 9-11h | 4373 (39 %) | 2867 (29 %) |  |
| 8-9h | 4630 (41 %) | 4413 (45 %) |  |
| 7-8h | 1659 (15 %) | 1820 (18 %) |  |
| 5-7h | 473 (4 %) | 680 (7 %) |  |
| <5h | 47 (0 %) | 60 (1 %) |  |
| **Chronotype** |  |  |  |
|  |  |  | <0.001 |
| Morning | 5907 (53 %) | 4371 (44 %) |  |
| Evening | 105 (1 %) | 123 (1 %) |  |
| Intermediate | 3191 (29 %) | 3807 (39 %) |  |
| Missing | 1979 (17.7%) | 1539 (15.6%) |  |
| **Social Jetlag** |  |  | <0.001 |
| mean (95% CI) | 2.01 (1.98, 2.04) | 2.19 (2.16, 2.23) |  |
| Missing | 515 (4.6%) | 8 (0.1%) |  |
| **Physical Activity** |  |  | <0.001 |
| mean (95% CI) | 3.51 (3.47, 3.55) | 3.71 (3.67, 3.75) |  |
| Missing | 19 (0.2%) | 11 (0.1%) |  |
| **Screen Time (hours)** |  |  | <0.001 |
| mean (95% CI) | 8.05 (7.94, 8.15) | 8.79 (8.69, 8.9) |  |
| Missing | 592 (5.3%) | 8 (0.1%) |  |
| **Social Responsiveness** |  |  | <0.001 |
| mean (95% CI) | 14.39 (14.31, 14.46) | 14.33 (14.25, 14.41) |  |
| **Externalising Symptoms** |  |  | <0.001 |
| mean (95% CI) | 45.21 (45.02, 45.39) | 44.3 (44.11, 44.48) |  |
| Missing | 2 (0.0%) | 5 (0.1%) |  |
| **Internalising Symptomst** |  |  | <0.001 |
| mean (95% CI) | 48.59 (48.39, 48.79) | 47.74 (47.53, 47.95) |  |
| Missing | 2 (0.0%) | 5 (0.1%) |  |
| **Cognition** |  |  |  |
| mean (95% CI) | 47.85 (47.64, 48.07) |  |  |
| Missing | 914 (8.2%) |  |  |

**Table 2.** Hierarchical mixed-effects regression model examining associations between total sleep disruption score and social responsiveness scores, adjusted for chronotype and demographic covariables

| *Predictors* | **Model 1** |  |  | **Model 2** |  |  | **Model 3** |  |  |
| --- | --- | --- | --- | --- | --- | --- | --- | --- | --- |
|  | *Estimates* | *CI* | *p* | *Estimates* | *CI* | *p* | *Estimates* | *CI* | *p* |
| Social Responsiveness | 0.58 | 0.55 – 0.61 | **<0.001** | 0.57 | 0.53 – 0.62 | **<0.001** | 0.12 | 0.07 – 0.17 | **<0.001** |
| Sex [ref = male] | 0.61 | 0.35 – 0.86 | **<0.001** | 0.62 | 0.25 – 0.99 | **0.001** | 0.67 | 0.33 – 1.01 | **<0.001** |
| Age [ref = 11 years old] | -0.62 | -0.76 – -0.48 | **<0.001** | -0.84 | -1.02 – -0.65 | **<0.001** | -0.51 | -0.70 – -0.32 | **<0.001** |
| Chronotype [Evening] |  |  |  | 1.43 | 0.24 – 2.61 | **0.018** | 2.17 | 1.03 – 3.32 | **<0.001** |
| Chronotype [Intermediate] |  |  |  | 1.09 | 0.80 – 1.37 | **<0.001** | 0.99 | 0.72 – 1.27 | **<0.001** |
| Physical Activity (hr/day) |  |  |  | 0.07 | 0.01 – 0.12 | **0.019** | 0.07 | 0.02 – 0.13 | **0.007** |
| Screen Time (hr/day) |  |  |  | 0.09 | 0.06 – 0.12 | **<0.001** | 0.05 | 0.03 – 0.08 | **<0.001** |
| Externalising Symptoms |  |  |  |  |  |  | 0.16 | 0.14 – 0.17 | **<0.001** |
| Internalising Symptoms |  |  |  |  |  |  | 0.21 | 0.19 – 0.22 | **<0.001** |
| Cognition |  |  |  |  |  |  | 0.02 | 0.01 – 0.03 | **0.004** |
| SRS × chronotype [evening] |  |  |  |  |  |  | 0.32 | 0.02 – 0.61 | **0.035** |
| SRS × chronotype [Intermediate]] |  |  |  |  |  |  | 0.18 | 0.11 – 0.25 | **<0.001** |
| SRS × sex [female] |  |  |  |  |  |  | 0.12 | 0.03 – 0.21 | **0.009** |

Additionally adjusted for SEP, pubertal stage, study site, ethnicity

Sequential hierarchical models were fitted, with Model 1 minimally adjusted for sex, age, race/ethnicity, and assessment wave, Model 2 additionally adjusted for data collection site, socioeconomic position, pubertal stage, physical activity, recreational screen use and chronotype. An interaction term between SRS and chronotype, and SRS and sex was included to test for differential associations by chronotype subgroup or sex. The final fully adjusted model (Model 3) further controlled for behavioural comorbidities, specifically internalizing and externalizing symptoms derived from the Child Behaviour Checklist. All models were estimated using maximum likelihood with a Gaussian distribution and identity link function. Model diagnostics confirmed homoscedasticity and approximate normality of residuals. Estimates are reported as unstandardized regression coefficients with corresponding standard errors and 95% confidence intervals. Collinearity was low with variance inflation factors less than 3 for all variables included in the models. The association of sleep disturbance among siblings was assessed using a likelihood ratio test to compare the fully adjusted model with a model with no random intercept for family. An intraclass correlation coefficient (ICC) was estimated from the fully adjusted mixed model as a descriptive measure of the variance explained by family membership (sibling random effect); 95% confidence intervals for the ICC were obtained from 500 parametric bootstrap replicates. Statistical significance was accepted at p < 0.05 and all analyses were performed in R version 4.5.1.

## Results

Sociodemographic parameters are shown as descriptive data of the sample of ABCD Study participants included in this study (Table 1) at the 1-year (n = 11,205; age 11 years) and 3-year (n = 10,108; age 13 years) follow-up waves. The sample was balanced by sex across waves (52% male) and the majority of participants identified as White (∼70%). Sleep duration was lower, chronotype later, social jetlag higher, and sleep disturbance score lower at 13 years compared to 11 years (Table 1). In terms of lifestyle parameters, screen time and physical activity were higher in mid- compared to early-adolescence. SRS was marginally lower at 13 years compared to 11 years, as were signs of internalising and externalising symptoms (Table 1).

A series of hierarchical mixed-effects regression models were applied to test the association between SRS and sleep disturbance, adjusting for chronotype and covariables (Figure 1A; Table 2). In the fully adjusted model (Model 3), higher SRS was significantly associated with greater sleep disturbance (β = 0.12, 95% CI 0.07–0.17; p < 0.001). Chronotype was also associated with sleep, with evening types (β = 2.17, 95% CI 1.03–3.32; p < 0.001) and intermediate types (β = 0.99, 95% CI 0.72–1.27; p < 0.001) reporting more sleep disturbance compared to morning types. There were significant interactions between SRS and chronotype, indicating that the association between social difficulties and sleep disturbance was stronger among evening types (β = 0.32, 95% CI 0.02–0.61; p = 0.035) and intermediate types (β = 0.18, 95% CI 0.11–0.25; p < 0.001), relative to morning types (Figure 1B). The final model accounted for 27.3% of the variance in sleep disturbance (marginal R² = 0.273) (Table 4, S5). Inclusion of a sibling-level random effect significantly improved model fit compared to a model with only individual-level random effects (LRT χ² = 119.68, df = 1, p < 0.001), indicating significant clustering of sleep disturbance among siblings. The ICC for sibling similarity was 0.37 (95% CI: 0.32–0.42) indicating correlation of sleep disturbance outcomes among siblings after adjustment for covariables.

**Figure 1:**
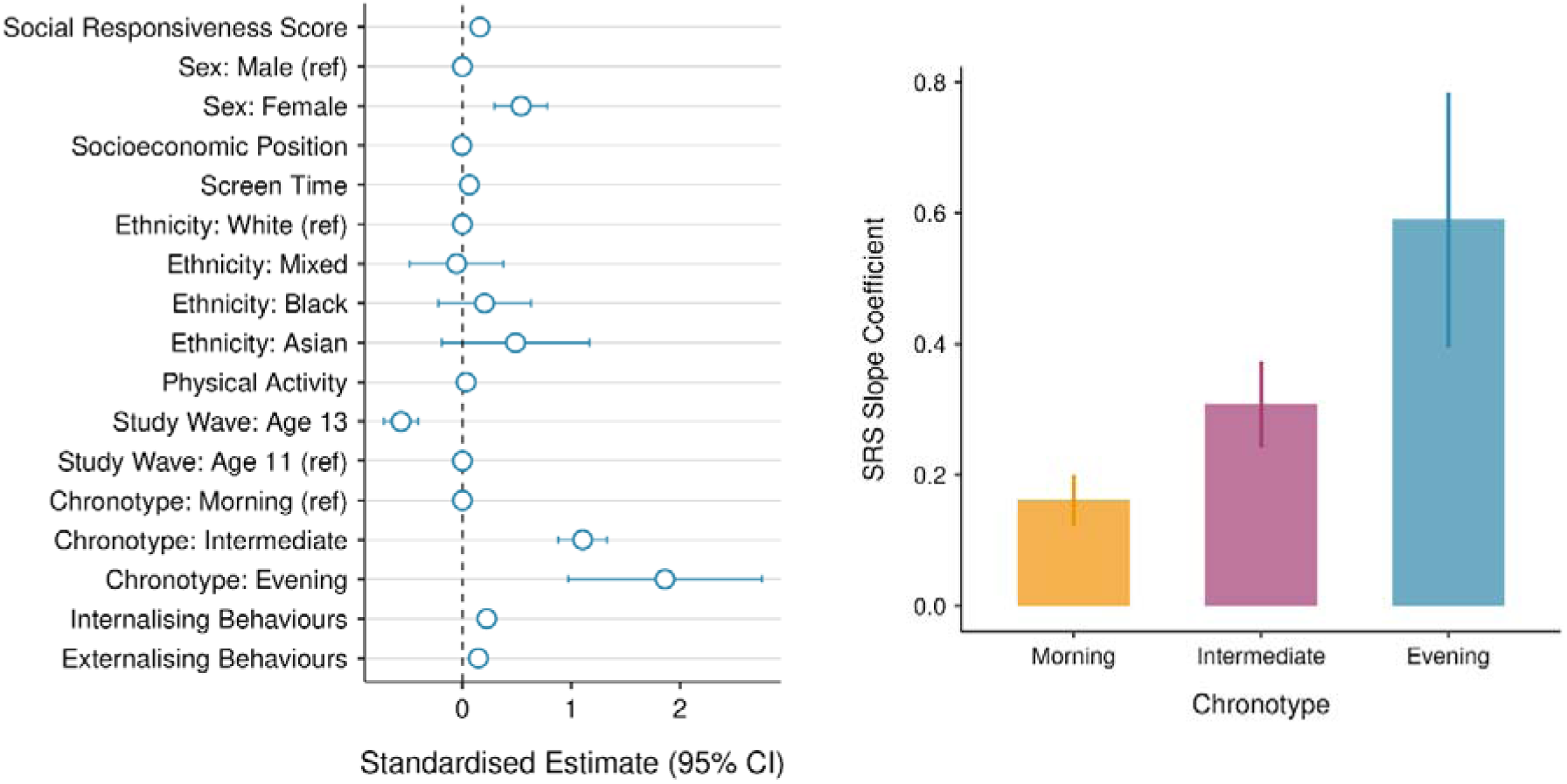
(A) Forest plot showing standardized beta estimates and 95% confidence intervals for the association between SRS and sleep disruption. Reference categories are coded with an estimated value of zero and the dashed vertical line denotes the null value (β = 0). (B) Estimated slopes and 95% confidence intervals for the association between SRS and sleep disruption, by chronotype. Estimates reflect the sum of the main SRS effect and the respective SRS × Chronotype interaction term.

A second hierarchical mixed-effects model tested the association between SRS and social jetlag (Table S5). In the fully adjusted model, SRS was not associated with social jetlag (β = –0.00, 95% CI –0.01–0.00, p = 0.733). Chronotype, however, was strongly related: evening types showed substantially greater social jetlag (β = 5.09, 95% CI 4.92–5.25, p < 0.001) and intermediate types also showed higher social jetlag (β = 1.51, 95% CI 1.47–1.55, p < 0.001), compared to morning types. Short sleep duration was similarly associated with greater social jetlag (< 5h: β = 0.63, 95% CI 0.35–0.90, p < 0.001, relative to 9–11h) (Table S6).

## Discussion

The results of this study demonstrate that social responsiveness was associated with sleep disturbance at a population-level, and that this relationship persisted across early adolescence and was modulated by chronotype independent of other behavioural symptoms or demographic and lifestyle factors. Furthermore, sleep disturbance was correlated among siblings, independent of individual-level random effects and covariates. These results of the largest investigation to date of social responsiveness and sleep disturbance illustrate the significance of sleep problems in children with autistic traits and is further evidence for an important role of sleep and circadian timing in autism.

Children with an autism diagnosis at baseline were excluded from the ABCD Study, so it was not possible to perform case-control studies in this dataset. However, autistic traits and their underlying neurobiological correlates exist in a continuum that extends into the general population^39,40^ and we used the SRS results as a proxy measure of the autistic phenotype in the ABCD Study. The SRS is not specifically associated with autistic traits, but we accounted for comorbid psychopathology in all of our models and previous studies have related SRS scores with the severity of behavioural signs and with subcortical pathology consistent with autism.^39,40^

Our findings in the ABCD Study corroborate previous studies that assessed the association between autistic traits and sleep disturbance in both case control and population-based studies and at different timepoints through childhood and adolescence. Autistic diagnosis and traits were associated with shorter sleep duration, but not with sleep efficiency in a sample of 157 17 year-old neurotypical children,^41^ and with bedtime resistance and increased sleep duration in 1,310 2-7 year old autistic and typically developing children.^3^ In a case-control study of 1,268 participants of the *Australian Autism Biobank*, children with autism were more likely to have sleep problems compared to their siblings or controls, and poor sleep was related to the presence and severity of social affect symptoms, restrictive behaviours and sensory problems.^1^ Similarly, reduced sleep duration was directly related to the severity of autistic symptoms in another child autism cohort, the *Simons Simplex Collection*.^14^ In a prospective study of neurotypical children at risk of autism (with a first degree relative that is autistic), sleep disturbance was higher in those later diagnosed with autism from age 14 months.^12^ Sleep was associated with the dimensions of autistic behaviour and cognitive ability but significantly, this was not consistent over the study timepoints (5, 10 and 14 months), suggesting that the strength of association between sleep disturbance and autistic behaviours might vary across neurodevelopment^12^ as it does in animal models.^26^

In population level studies, sleep disturbance was associated with autistic traits in 5,151 children at all timepoints (1.5, 3, 6, 9 years old) assessed in the *Generation R Cohort*,^42^ from 30 months in the *ALSPC Study*,^8^ from 6 months in the *Borwon Infant Study,* ^43^ 12 months in *the Conditions Affecting Neurocognitive Development in Early Childhood (CANDLE) Study*^13^ and at just 1 month of age in the *Japan Environmental and Children Study*.^44^ In the *Bergen Child Study* (n = 3,700), sleep disturbance was related to autistic traits at both 7-9 and 13-15 years of age, with severity increasing over time only in those children above the threshold value for autism diagnosis.^1,10^ It is clear from both case-control studies and big cohort datasets that sleep disturbance is evident in early infancy and probably from birth in children later diagnosed with autism, and the association is sustained and proportional to the severity of behavioural signs. This is consistent with our findings of a longitudinal and independent association between autistic traits and sleep disturbance across early adolescence in the participants of the ABCD Study. Sleep disturbance in infancy is probably the first indicator of autism, that precedes and is at the very least, closely associated with future behavioural symptoms.

In this study, the association between sleep disturbance and autistic traits was sex dependent, with girls with high SRS scores more likely to have sleep disturbance compared to boys. The association between sleep and autistic-like behaviours was sex-dependent in animal models,^24^ and in autistic children^1^ and adults,^45^ where females were found to have worse autism-related sleep problems compared to males. This is consistent with the different behavioural presentations of autism between male and females, as well as with a general increased prevalence of sleep problems in women. Sleep disturbance was correlated among siblings in the general population sample that the ABCD Study represents, reflecting familial aggregation independent of individual-level covariates.

Childhood and adolescence, and in particular the first year of life, are critical periods of synaptic remodelling and the development of sensory and cortical networks^27,46,47^ during which the brain is profoundly vulnerable to the effects of sleep disturbance. Animals are more susceptible to the effects of sleep disturbance in early life; mice in the pre-adolescent phase of development (P21-28) completely lack the adaptive responses to sleep deprivation of adults (P70-100) (increased sleep latency, rebound active phase recovery, *Homer1a* expression, increased slow wave activity) and instead show reduced memory performance, and upregulation of synaptic proteins including some encoded by autism risk genes.^26^ Sleep disturbance in adolescent (P35-42) but not adult mice, had enduring effects on social novelty preference that were rescued by improving NREM sleep.^27^ The profound effects of sleep disturbance in early life on social behaviour in animal models are consistent with the independent, cross-sectional and prospective associations between sleep disturbance and indicators of autism in adolescence reported in this study and others.

In the present study using data from the ABCD Study, we found that the association between autistic traits and sleep disturbance was modulated by chronotype, with a stronger association in children with evening chronotypes. Chronotype is an output of the human circadian clock and its modulatory effect in this study further supports a regulatory role for circadian mechanisms in the relationship between sleep disturbance and autistic traits. Our findings are in partial agreement with a small case-control study that reported that evening chronotype and social jetlag correlated with psychiatric outcomes in 12-year-old autistic children,^6^ although social jetlag was not associated with autistic traits in our study. In contrast, previous investigations of the ABCD Study reported that social jetlag was associated with poor cognitive development and altered cortical functional connectivity,^48,49^ although social responsiveness was not addressed. Studies in adults report complex relationships between autism, sleep and chronotype; autism was associated with evening chronotype as a main effect, but sleep disturbance was independent of chronotype in two small case-control studies in autistic adults.^45,50^ Our findings of strong associations of autistic traits with sleep disturbance but not social jetlag in early adolescence, as well as discrepancies with studies in autistic adults where chronotype was independent of sleep disturbance,^45,50^ suggest that associations between sleep disturbance and autistic behaviours might depend on the stage of neurodevelopment. This aligns with finding in animal models where the behavioural effects of sleep and circadian disruption are strongly dependent on the stage of development^24,26,27^ and the nature of the disruption. For example, juvenile mice (P21-28) were more vulnerable to the effects of sleep deprivation on behaviour, while adolescents (P42-49) were more susceptible to the effects of circadian disruption.^26^

The timing of critical periods of postnatal cortical development is regulated by the circadian clock^46^ and a core defect in the circadian timing mechanism could cause sleep disturbance and alter critical period timing that together deviate the trajectory of brain development^15–18^ and contribute to the life-long changes in social behaviour and sleep disturbance that characterise autism. Neurobiological changes in animal models of autism may not represent the human condition, but their parallels with epidemiological associations such as those reported here, must be considered in the absence of reliable methods for mechanistic investigations in humans. Importantly, methods that do permit causal inference (genetic correlation, Mendelian Randomisation studies) support a role for sleep in the aetiology of autism and corroborate the findings of animal models.^28,29^ Collectively, the evidence supports a mechanistic model whereby circadian disruption during critical windows of neurodevelopment interacts with underlying genetic liability to drive sex-specific alterations in social behaviour and sleep problems that persist into adulthood.

The results and conclusions of this study must be considered in context of several serious limitations. The ABCD Study excluded children with autism or intellectual disability at recruitment, so the autistic traits reported in this study do not span the range of behaviour of autistic children, and our results may not be comparable to studies without this restriction. Conversely, this might also be a strength, since in clinical populations sleep disturbance could occur secondary to more severe autistic behaviours and not exclusively as a feature of the condition. Our studies assume that autism exists as a continuous trait through the population which might not be justified, and sleep disturbance in people with subclinical autistic traits might differ from that of people with an autism diagnosis. Deficits in social responsiveness are not exclusive to autism and comorbid neurodevelopmental conditions such as ADHD or OCD could affect the results of the SRS. The large sample size of the ABCD Study is a strength of this study, as is the detailed demographic and lifestyle information that allowed adjustment for multiple lifestyle factors and behavioural comorbidities. The observational and caregiver-reported sleep and behavioural data is subject to bias and our results require reproduction in datasets where objective sleep data are available. The present study was limited to analysis of sleep disturbance and autistic traits at only two timepoints (ages 11 and 13 years), but the ABCD Study will eventually contain data at timepoints that span adolescence and that will allow investigation of the developmental trajectories of sleep and circadian disruption, and their relative association with autistic traits.

In this study, autistic traits assessed in early adolescence were associated with sleep disturbance independent of behavioural comorbidities and multiple confounding factors and prospectively associated with sleep disturbance at age 13 years. Manipulation of sleep stages rescued the effects of sleep disturbance on autistic behaviours in animals,^27^ and future research should investigate whether interventions that regulate circadian rhythms from birth can ameliorate the effects of disrupted sleep on neurodevelopment in children susceptible to autism. Research to develop objective, low-burden and ecologically valid methods for longitudinal monitoring sleep from birth is urgently required to further understanding of how sleep and circadian rhythms shape neurodevelopment. The close associations between sleep disturbance and autistic traits across early adolescence in this study add to the evidence implicating disrupted sleep and circadian timing as endophenotypes of autism that connect the genetic risk of this condition to the behavioural phenotype.

## Data Availability

Data are available to registered researchers online at https://abcdstudy.org/

## Data sharing

Data are from ABCD data release 5.0.0 (released in June 2023 DOI: 10.15154/8873-zj65; a related data dictionary can be found at https://data-dict.abcdstudy.org/).

