## supplement for "Associations Between Social Responsiveness and Sleep disturbance are Modulated by Chronotype in Early Adolescence: Cross-Sectional and Prospective Findings from 10,108 Participants of the Adolescent Brain and Cognitive Development (ABCD) Study"

Table S1	Details of the questions administered in the SRS

| Table Name | Name | Variable Label | Notes | Table Name (NDA) |
| --- | --- | --- | --- | --- |
| mh_p_ssrs | ssrs_6_p | Would rather be alone than with others. | 1 = Not True No  2 = Sometimes True  3 = Often True  4 = Almost Always True | abcd_pssrs01 |
| mh_p_ssrs | ssrs_15r_p | Is able to understand the meaning of other people's tone of voice and facial expressions. | 4 = Not True No  3 = Sometimes True  2 = Often True  1 = Almost Always True | abcd_pssrs01 |
| mh_p_ssrs | ssrs_16_p | Avoids eye contact or has unusual eye contact | 1 = Not True No  2 = Sometimes True;  3 = Often True  4 = Almost Always True | abcd_pssrs01 |
| mh_p_ssrs | ssrs_18_p | Has difficulty making friends, even when trying his or her best. | 1 = Not True No  2 = Sometimes True  3 = Often True  4 = Almost Always True | abcd_pssrs01 |
| mh_p_ssrs | ssrs_24_p | Has more difficulty than other children with changes in his or her routine. | 1 = Not True No  2 = Sometimes  3 = Often True  4 = Almost Always True | abcd_pssrs01 |
| mh_p_ssrs | ssrs_29_p | Is regarded by other children as odd or weird. | 1 = Not True No  2 = Sometimes True  3 = Often True  4 = Almost Always True | abcd_pssrs01 |
| mh_p_ssrs | ssrs_35_p | Has trouble keeping up with the flow of normal conversation. | 1 = Not True No  2 = Sometimes True  3 = Often True  4 = Almost Always True | abcd_pssrs01 |
| mh_p_ssrs | ssrs_37_p | Has difficulty relating to peers. | 1 = Not True No  2 = Sometimes True  3 = Often True  4 = Almost Always True | abcd_pssrs01 |
| mh_p_ssrs | ssrs_39_p | Has an unusually narrow range of interests. | 1 = Not True No  2 = Sometimes True  3 = Often True  4 = Almost Always True | abcd_pssrs01 |
| mh_p_ssrs | ssrs_42_p | Seems overly sensitive to sounds, textures or smells. | 1 = Not True No  2 = Sometimes True  3 = Often True  4 = Almost Always True | abcd_pssrs01 |
| mh_p_ssrs | ssrs_58_p | Concentrates too much on parts of things rather than seeing the whole picture. | 1 = Not True No  2 = Sometimes True  3 = Often True  4 = Almost Always True | abcd_pssrs01 |
| mh_p_ssrs | ssrs_p_ss_sum | SSRS sum[ssrs_15r, ssrs_6, ssrs_16, ssrs_18, ssrs_24, ssrs_29, ssrs_35, ssrs_37, ssrs_39, ssrs_42, ssrs_58] |  | abcd_mhp02 |

Table S2	Questionnaires administered at each follow up wave.

| **Measure** | **Category** | **Subcategory** | **Source** | **Baseline** | **Follow Up Year** | | | | | |
| --- | --- | --- | --- | --- | --- | --- | --- | --- | --- | --- |
|  |  |  |  |  | **1** | **2** | **3** | **4** | **5** | **6** |
| SRS | Mental Health | Autism Spectrum | Parent | Yes | No | Yes | No | Yes | No | Yes |
| Munich Chronotype Questionnaire (sleep) | Physical Health | Sleep | Youth | No | No | Yes | Yes | Yes | Yes | Yes |
| Child Behavior Checklist | Mental Health | Broad Psychopathology | Parent | Yes | Yes | Yes | Yes | Yes | Yes | Yes |

Table S3	Details of the timepoints used for Time 1 and Time 2 for each variable included in the dataset in this study. Some variables were imputed from the nearest available time point to address missing data Note that SRS was only measured at Year 1.

| **Var Name** | **Time 1** | **Var Code in ABCD** | **Time 2** |
| --- | --- | --- | --- |
| SRS | 1_year_follow_up_y_arm_1 | ssrs_p_ss_sum | 1_year_follow_up_y_arm_1 |
| CBCL_ext | 1_year_follow_up_y_arm_1 | cbcl_scr_syn_external_t | 3_year_follow_up_y_arm_1 |
| CBCL_int | 1_year_follow_up_y_arm_1 | cbcl_scr_syn_internal_t | 3_year_follow_up_y_arm_1 |
| Chronotype | 2_year_follow_up_y_arm_1 | mctq_msfsc_correct | 4_year_follow_up_y_arm_1 |
| SJL | 2_year_follow_up_y_arm_1 | mctq_sjlrel_calc | 4_year_follow_up_y_arm_1 |
| Sleep | 1_year_follow_up_y_arm_1 | Sleep Disturbance | 3_year_follow_up_y_arm_1 |
| PA | 1_year_follow_up_y_arm_1 | physical_activity1_y | 3_year_follow_up_y_arm_1 |
| Cognition | baseline_year_1_arm_1 | nihtbx_totalcomp_fc | baseline_year_1_arm_1 |
| Screens | 2_year_follow_up_y_arm_1 | screentime_total_p | 3_year_follow_up_y_arm_1 |
| Interview Age | 1_year_follow_up_y_arm_1 | interview_age | 3_year_follow_up_y_arm_1 |
| Sex | baseline_year_1_arm_1 | sex | baseline_year_1_arm_1 |
| Ethnicity | 1_year_follow_up_y_arm_1 | race.4level.p | baseline_year_1_arm_1 |
| SEP | baseline_year_1_arm_1 | reshist_addr1_adi_perc | baseline_year_1_arm_1 |
| Income | 1_year_follow_up_y_arm_1 | household.income.3level |  |
| Site | 1_year_follow_up_y_arm_1 | site_id_l | 3_year_follow_up_y_arm_1 |

Table S4	Demographic, lifestyle and behavioural characteristics between children classified as having normal sleep (n = 15,615) and those with sleep disturbance (n = 5,425) at baseline (Follow-up Year 1, age 11).

|  | **Normal Sleep (N=8170)** | **Sleep Disturbance (N=3012)** | **p** |
| --- | --- | --- | --- |
| **Sex** |  |  | NS |
| male | 4253 (52 %) | 1571 (52 %) |  |
| female | 3912 (48 %) | 1440 (48 %) |  |
| Missing | 5 (0.1%) | 1 (0.0%) |  |
| **Age (baseline)** |  |  | NS |
| mean (95% CI) | 131.08 (130.91, 131.24) | 131.09 (130.82, 131.37) |  |
| Missing | 1 (0.0%) | 0 (0%) |  |
| **Puberty** |  |  | NS |
| early puberty | 1362 (17 %) | 428 (14 %) |  |
| mid puberty | 1805 (22 %) | 656 (22 %) |  |
| late puberty | 1428 (17 %) | 498 (17 %) |  |
| post puberty | 789 (10 %) | 326 (11 %) |  |
| Missing | 2786 (34.1%) | 1104 (36.7%) |  |
| **Ethnicity** |  |  | NS |
| White | 5739 (70 %) | 2103 (70 %) |  |
| Black | 1183 (14 %) | 426 (14 %) |  |
| Asian | 287 (4 %) | 104 (3 %) |  |
| Other/Mixed | 764 (9 %) | 322 (11 %) |  |
| Missing | 197 (2.4%) | 57 (1.9%) |  |
| **SEP** |  |  | **<0.001** |
| mean (95% CI) | 38.97 (38.37, 39.57) | 41.53 (40.52, 42.55) |  |
| Missing | 591 (7.2%) | 204 (6.8%) |  |
| **Sleep Disturbance** |  |  | **<0.001** |
| 9-11h | 3568 (44 %) | 805 (27 %) |  |
| 8-9h | 3419 (42 %) | 1211 (40 %) |  |
| 7-8h | 1028 (13 %) | 631 (21 %) |  |
| 5-7h | 145 (2 %) | 328 (11 %) |  |
| <5h | 10 (0 %) | 37 (1 %) |  |
| **Chronotype** |  |  | **<0.001** |
| Morning | 4501 (55 %) | 1406 (47 %) |  |
| Evening | 64 (1 %) | 41 (1 %) |  |
| Intermediate | 2195 (27 %) | 996 (33 %) |  |
| Missing | 1410 (17.3%) | 569 (18.9%) |  |
| **Social Jetlag** |  |  | **<0.001** |
| mean (95% CI) | 1.94 (1.9, 1.97) | 2.21 (2.15, 2.28) |  |
| Missing | 360 (4.4%) | 155 (5.1%) |  |
| **Physical Activity** |  |  | **<0.001** |
| mean (95% CI) | 3.52 (3.47, 3.57) | 3.48 (3.4, 3.57) |  |
| Missing | 13 (0.2%) | 6 (0.2%) |  |
| **Screens** |  |  | **<0.001** |
| mean (95% CI) | 7.74 (7.62, 7.86) | 8.88 (8.66, 9.1) |  |
| Missing | 409 (5.0%) | 183 (6.1%) |  |
| **Social Responsiveness** |  |  |  |
| mean (95% CI) | 13.63 (13.56, 13.7) | 16.43 (16.24, 16.62) | **<0.001** |
| **Externalising Symptoms** |  |  | **<0.001** |
| mean (95% CI) | 43.03 (42.83, 43.22) | 51.11 (50.73, 51.5) |  |
| Missing | 2 (0.0%) | 0 (0%) |  |
| **Internalising Symptoms** |  |  | **<0.001** |
| mean (95% CI) | 45.95 (45.75, 46.15) | 55.74 (55.36, 56.12) |  |
| Missing | 2 (0.0%) | 0 (0%) |  |
| **Cognition** |  |  | **<0.001** |
| mean (95% CI) | 48.02 (47.77, 48.27) | 47.41 (46.98, 47.84) |  |
| Missing | 665 (8.1%) | 249 (8.3%) |  |

Table S5	Hierarchical mixed-effects regression model examining associations between total sleep disruption score and social responsiveness scores, adjusted for chronotype and demographic covariables

|  |  | | | **Sleep Disturbance** | | |  | | |
| --- | --- | --- | --- | --- | --- | --- | --- | --- | --- |
| *Predictors* | *Estimates* | *CI* | *p* | *Estimates* | *CI* | *p* | *Estimates* | *CI* | *p* |
| **Social Responsiveness** | 0.58 | 0.55 – 0.61 | **<0.001** | 0.57 | 0.53 – 0.62 | **<0.001** | 0.12 | 0.07 – 0.17 | **<0.001** |
| **Sex** [female] | 0.61 | 0.35 – 0.86 | **<0.001** | 0.62 | 0.25 – 0.99 | **0.001** | 0.67 | 0.33 – 1.01 | **<0.001** |
| **Ethnicity** [Black] | -0.06 | -0.45 – 0.33 | 0.754 | -0.99 | -1.68 – -0.31 | **0.004** | -0.20 | -0.83 – 0.43 | 0.529 |
| **Ethnicity** [Asian] | -0.59 | -1.32 – 0.13 | 0.108 | -0.11 | -1.09 – 0.87 | 0.822 | 0.80 | -0.09 – 1.68 | 0.077 |
| **Ethnicity** [Other/Mixed] | 0.02 | -0.43 – 0.47 | 0.936 | 0.10 | -0.52 – 0.73 | 0.744 | -0.09 | -0.65 – 0.47 | 0.756 |
| **Study Timepoint** [13 years old] | -0.62 | -0.76 – -0.48 | **<0.001** | -0.84 | -1.02 – -0.65 | **<0.001** | -0.51 | -0.70 – -0.32 | **<0.001** |
| **ABCD Study Site** [site02] |  |  |  | 0.05 | -1.26 – 1.35 | 0.944 | -0.12 | -1.30 – 1.07 | 0.846 |
| **ABCD Study Site** [site03] |  |  |  | -0.63 | -1.95 – 0.68 | 0.345 | -0.71 | -1.92 – 0.51 | 0.255 |
| **ABCD Study Site** [site04] |  |  |  | 1.21 | -0.07 – 2.48 | 0.064 | 0.89 | -0.28 – 2.05 | 0.137 |
| **ABCD Study Site** [site05] |  |  |  | 1.28 | -0.14 – 2.71 | 0.076 | 0.88 | -0.41 – 2.18 | 0.180 |
| **ABCD Study Site** [site06] |  |  |  | 0.75 | -0.50 – 1.99 | 0.238 | 0.33 | -0.82 – 1.48 | 0.572 |
| **ABCD Study Site** [site07] |  |  |  | -0.63 | -2.15 – 0.89 | 0.418 | -0.34 | -1.73 – 1.04 | 0.626 |
| **ABCD Study Site** [site08] |  |  |  | 0.34 | -1.07 – 1.74 | 0.637 | -0.24 | -1.54 – 1.06 | 0.718 |
| **ABCD Study Site** [site09] |  |  |  | -0.77 | -2.12 – 0.58 | 0.266 | -0.54 | -1.77 – 0.70 | 0.397 |
| **ABCD Study Site** [site10] |  |  |  | -1.15 | -2.39 – 0.09 | 0.068 | -1.08 | -2.22 – 0.05 | 0.061 |
| **ABCD Study Site** [site11] |  |  |  | 0.88 | -0.56 – 2.32 | 0.233 | 0.71 | -0.60 – 2.02 | 0.286 |
| **ABCD Study Site** [site12] |  |  |  | 0.69 | -0.66 – 2.03 | 0.318 | 0.31 | -0.92 – 1.53 | 0.623 |
| **ABCD Study Site** [site13] |  |  |  | 0.19 | -1.08 – 1.46 | 0.769 | -0.36 | -1.55 – 0.82 | 0.546 |
| **ABCD Study Site** [site14] |  |  |  | -1.07 | -2.37 – 0.23 | 0.106 | -0.61 | -1.80 – 0.58 | 0.314 |
| **ABCD Study Site** [site15] |  |  |  | 1.00 | -0.53 – 2.54 | 0.199 | 0.93 | -0.52 – 2.38 | 0.211 |
| **ABCD Study Site** [site16] |  |  |  | 0.22 | -0.97 – 1.40 | 0.720 | -0.47 | -1.55 – 0.62 | 0.398 |
| **ABCD Study Site** [site17] |  |  |  | -0.13 | -1.40 – 1.14 | 0.840 | -0.65 | -1.83 – 0.53 | 0.281 |
| **ABCD Study Site** [site18] |  |  |  | 0.81 | -0.55 – 2.18 | 0.243 | 0.32 | -0.92 – 1.56 | 0.608 |
| **ABCD Study Site** [site19] |  |  |  | -1.15 | -2.62 – 0.33 | 0.128 | -1.04 | -2.41 – 0.32 | 0.135 |
| **ABCD Study Site** [site20] |  |  |  | -0.97 | -2.27 – 0.33 | 0.144 | -0.95 | -2.13 – 0.23 | 0.116 |
| **ABCD Study Site** [site21] |  |  |  | -0.09 | -1.39 – 1.21 | 0.897 | -0.34 | -1.54 – 0.86 | 0.579 |
| **Puberty** [mid puberty] |  |  |  | 0.16 | -0.26 – 0.58 | 0.448 | -0.01 | -0.40 – 0.37 | 0.943 |
| **Puberty** [late puberty] |  |  |  | 0.12 | -0.33 – 0.57 | 0.604 | 0.05 | -0.37 – 0.46 | 0.829 |
| **Puberty** [post puberty] |  |  |  | 0.85 | 0.30 – 1.40 | **0.002** | 0.60 | 0.10 – 1.11 | **0.019** |
| **SEP** |  |  |  | 0.00 | -0.01 – 0.01 | 0.426 | -0.00 | -0.01 – 0.01 | 0.773 |
| **Chronotype** [Evening] |  |  |  | 1.43 | 0.24 – 2.61 | **0.018** | 2.17 | 1.03 – 3.32 | **<0.001** |
| **Chronotype** [Intermediate] |  |  |  | 1.09 | 0.80 – 1.37 | **<0.001** | 0.99 | 0.72 – 1.27 | **<0.001** |
| **Physical Activity** |  |  |  | 0.07 | 0.01 – 0.12 | **0.019** | 0.07 | 0.02 – 0.13 | **0.007** |
| **Screen Time** |  |  |  | 0.09 | 0.06 – 0.12 | **<0.001** | 0.05 | 0.03 – 0.08 | **<0.001** |
| **Externalising Symptoms** |  |  |  |  |  |  | 0.16 | 0.14 – 0.17 | **<0.001** |
| **Internalising Symptoms** |  |  |  |  |  |  | 0.21 | 0.19 – 0.22 | **<0.001** |
| **Cognition** |  |  |  |  |  |  | 0.02 | 0.01 – 0.03 | **0.004** |
| **Social Responsiveness** × **Chronotype** [Evening] |  |  |  |  |  |  | 0.32 | 0.02 – 0.61 | **0.035** |
| **Social Responsiveness × Chronotype** [Intermediate] |  |  |  |  |  |  | 0.18 | 0.11 – 0.25 | **<0.001** |
| **Social Responsiveness × Sex** [female] |  |  |  |  |  |  | 0.12 | 0.03 – 0.21 | **0.009** |
| **Random Effects** | | | | | | | | | |
| σ^2^ | 24.48 | | | 20.75 | | | 19.62 | | |
| τ_00_ | 9.35 _src_subject_id:related_ | | | 8.21 _src_subject_id:related_ | | | 6.24 _src_subject_id:related_ | | |
|  | 23.65 _related_ | | | 22.76 _related_ | | | 14.98 _related_ | | |
| ICC | 0.57 | | | 0.60 | | | 0.52 | | |
| N | 10940 _src_subject_id_ | | | 6189 _src_subject_id_ | | | 5718 _src_subject_id_ | | |
|  | 9058 _related_ | | | 5440 _related_ | | | 5077 _related_ | | |
| Observations | 20572 | | | 10737 | | | 9915 | | |
| Marginal R^2^ / Conditional R^2^ | 0.090 / 0.613 | | | 0.118 / 0.646 | | | 0.279 / 0.653 | | |

Table S6	Hierarchical mixed-effects regression model examining associations between social jetlag and social responsiveness scores, adjusted for chronotype and demographic covariables.

|  |  | | | **Social Jetlag** | | |  | | |
| --- | --- | --- | --- | --- | --- | --- | --- | --- | --- |
| *Predictors* | *Estimates* | *CI* | *p* | *Estimates* | *CI* | *p* | *Estimates* | *CI* | *p* |
| **Social Responsiveness** | 0.01 | 0.00 – 0.01 | **0.012** | 0.00 | -0.00 – 0.01 | 0.524 | -0.00 | -0.01 – 0.00 | 0.733 |
| **Sex** [female] | 0.11 | 0.05 – 0.16 | **<0.001** | 0.11 | 0.07 – 0.15 | **<0.001** | 0.11 | 0.07 – 0.15 | **<0.001** |
| **Ethnicity** [Black] | 1.39 | 1.31 – 1.47 | **<0.001** | 0.42 | 0.35 – 0.49 | **<0.001** | 0.42 | 0.34 – 0.49 | **<0.001** |
| **Ethnicity** [Asian] | -0.30 | -0.45 – -0.16 | **<0.001** | -0.11 | -0.23 – 0.00 | 0.052 | -0.11 | -0.23 – 0.01 | 0.067 |
| **Ethnicity** [Other/Mixed] | 0.54 | 0.45 – 0.63 | **<0.001** | 0.09 | 0.02 – 0.16 | **0.014** | 0.09 | 0.01 – 0.16 | **0.023** |
| **Study Timepoint** [Age 13] | 0.21 | 0.18 – 0.24 | **<0.001** | -0.01 | -0.04 – 0.02 | 0.493 | -0.01 | -0.04 – 0.03 | 0.707 |
| **ABCD Study Site** [site02] |  |  |  | -0.06 | -0.21 – 0.10 | 0.461 | -0.04 | -0.20 – 0.13 | 0.664 |
| **ABCD Study Site** [site03] |  |  |  | 0.04 | -0.12 – 0.19 | 0.638 | 0.06 | -0.10 – 0.22 | 0.487 |
| **ABCD Study Site** [site04] |  |  |  | -0.02 | -0.17 – 0.13 | 0.808 | 0.01 | -0.14 – 0.17 | 0.868 |
| **ABCD Study Site** [site05] |  |  |  | 0.10 | -0.07 – 0.26 | 0.241 | 0.15 | -0.02 – 0.32 | 0.094 |
| **ABCD Study Site** [site06] |  |  |  | -0.40 | -0.55 – -0.25 | **<0.001** | -0.38 | -0.54 – -0.23 | **<0.001** |
| **ABCD Study Site** [site07] |  |  |  | -0.37 | -0.55 – -0.18 | **<0.001** | -0.29 | -0.48 – -0.10 | **0.003** |
| **ABCD Study Site** [site08] |  |  |  | -0.30 | -0.47 – -0.13 | **<0.001** | -0.29 | -0.47 – -0.11 | **0.002** |
| **ABCD Study Site** [site09] |  |  |  | -0.14 | -0.29 – 0.02 | 0.097 | -0.10 | -0.27 – 0.07 | 0.240 |
| **ABCD Study Site** [site10] |  |  |  | -0.01 | -0.15 – 0.14 | 0.916 | 0.02 | -0.13 – 0.18 | 0.765 |
| **ABCD Study Site** [site11] |  |  |  | -0.08 | -0.25 – 0.08 | 0.326 | -0.04 | -0.21 – 0.13 | 0.646 |
| **ABCD Study Site** [site12] |  |  |  | -0.23 | -0.39 – -0.08 | **0.003** | -0.18 | -0.34 – -0.02 | **0.026** |
| **ABCD Study Site** [site13] |  |  |  | -0.22 | -0.37 – -0.07 | **0.004** | -0.20 | -0.36 – -0.04 | **0.013** |
| **ABCD Study Site** [site14] |  |  |  | -0.16 | -0.31 – -0.00 | **0.043** | -0.11 | -0.27 – 0.05 | 0.174 |
| **ABCD Study Site** [site15] |  |  |  | -0.29 | -0.46 – -0.11 | **0.001** | -0.26 | -0.45 – -0.08 | **0.005** |
| **ABCD Study Site** [site16] |  |  |  | -0.26 | -0.40 – -0.12 | **<0.001** | -0.22 | -0.37 – -0.07 | **0.003** |
| **ABCD Study Site** [site17] |  |  |  | -0.27 | -0.42 – -0.12 | **<0.001** | -0.26 | -0.42 – -0.10 | **0.002** |
| **ABCD Study Site** [site18] |  |  |  | -0.14 | -0.30 – 0.02 | 0.092 | -0.09 | -0.26 – 0.08 | 0.282 |
| **ABCD Study Site** [site19] |  |  |  | -0.01 | -0.18 – 0.16 | 0.918 | -0.06 | -0.25 – 0.12 | 0.495 |
| **ABCD Study Site** [site20] |  |  |  | -0.05 | -0.21 – 0.10 | 0.480 | -0.03 | -0.19 – 0.13 | 0.728 |
| **ABCD Study Site** [site21] |  |  |  | -0.13 | -0.29 – 0.02 | 0.088 | -0.10 | -0.26 – 0.06 | 0.215 |
| **SEP** |  |  |  | 0.01 | 0.01 – 0.01 | **<0.001** | 0.01 | 0.00 – 0.01 | **<0.001** |
| **Chronotype** [Evening] |  |  |  | 5.12 | 4.96 – 5.29 | **<0.001** | 5.09 | 4.92 – 5.25 | **<0.001** |
| **Chronotype** [Intermediate] |  |  |  | 1.52 | 1.48 – 1.56 | **<0.001** | 1.51 | 1.47 – 1.55 | **<0.001** |
| **PA** |  |  |  | 0.00 | -0.01 – 0.01 | 0.949 | -0.00 | -0.01 – 0.01 | 0.430 |
| **Screen Time** |  |  |  | 0.01 | 0.00 – 0.01 | **0.002** | 0.01 | 0.00 – 0.01 | **0.004** |
| **Sleep Duration** [8-9h] |  |  |  | 0.05 | 0.01 – 0.09 | **0.013** | 0.06 | 0.01 – 0.10 | **0.008** |
| **Sleep Duration** [7-8h] |  |  |  | 0.11 | 0.05 – 0.16 | **<0.001** | 0.11 | 0.05 – 0.17 | **<0.001** |
| **Sleep Duration** [5-7h] |  |  |  | 0.23 | 0.14 – 0.32 | **<0.001** | 0.23 | 0.13 – 0.32 | **<0.001** |
| **Sleep Duration** [<5h] |  |  |  | 0.56 | 0.30 – 0.82 | **<0.001** | 0.63 | 0.35 – 0.90 | **<0.001** |
| **Cognition** |  |  |  |  |  |  | -0.00 | -0.01 – -0.00 | **<0.001** |
| **Externalising Symptoms** |  |  |  |  |  |  | 0.01 | 0.00 – 0.01 | **<0.001** |
| **Internalising Symptoms** |  |  |  |  |  |  | -0.00 | -0.01 – -0.00 | **0.002** |
| **Random Effects** | | | | | | | | | |
| σ^2^ | 1.40 | | | 0.96 | | | 0.95 | | |
| τ_00_ | 0.30 _src_subject_id:related_ | | | 0.09 _src_subject_id:related_ | | | 0.08 _src_subject_id:related_ | | |
|  | 0.77 _related_ | | | 0.26 _related_ | | | 0.26 _related_ | | |
| ICC | 0.43 | | | 0.27 | | | 0.26 | | |
| N | 10576 _src_subject_id_ | | | 9276 _src_subject_id_ | | | 8539 _src_subject_id_ | | |
|  | 8761 _related_ | | | 7890 _related_ | | | 7347 _related_ | | |
| Observations | 20066 | | | 15933 | | | 14649 | | |
| Marginal R^2^ / Conditional R^2^ | 0.096 / 0.487 | | | 0.462 / 0.606 | | | 0.466 / 0.607 | | |

Figure S1	Relationship between sleep disturbance and quartiles of SRS at age 11 and 13 years

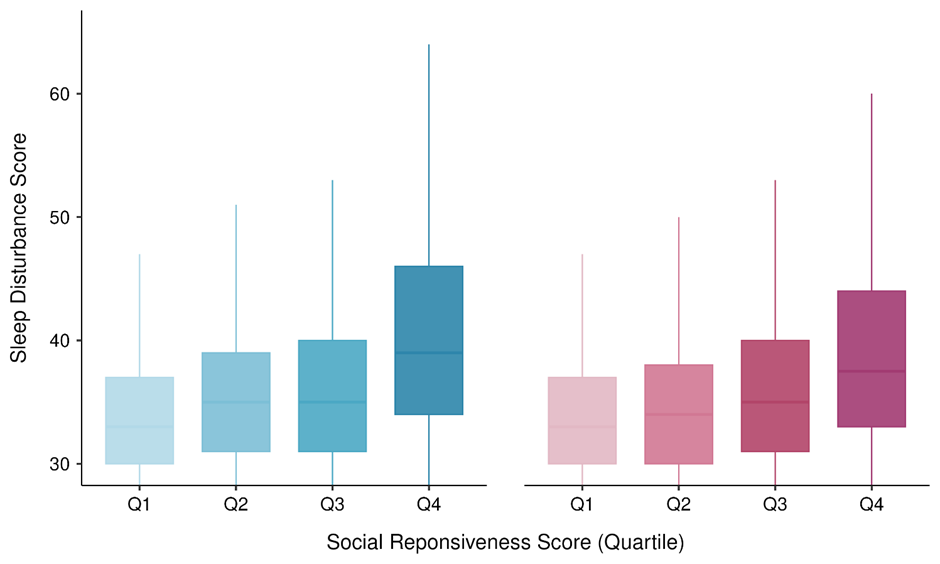

Age 11

Age 13
